# Ascorbic acid attenuates activation and cytokine production in sepsis-like monocytes

**DOI:** 10.1101/2021.04.15.21255504

**Authors:** Tobias Schmidt, Robin Kahn, Fredrik Kahn

## Abstract

Sepsis manifests due to the host’s dysregulated immune response to an infection. High dose ascorbic acid (AA) has emerged as a potential treatment of sepsis, yet little is known regarding how ascorbic acid influences the immune system in sepsis, such as monocytes. The objective of this study is to investigate the effects of high dose AA on monocyte polarization and cytokine production *in vitro*.

Monocytes were isolated from healthy donors (n=6) and polarized *in vitro* for 48hrs using lipopolysaccharide (LPS) and lipoteichoic acid (LTA). Polarization was confirmed by surface marker expression using flow cytometry. In parallel, monocytes from septic patients (n=3) were analyzed for polarization markers as a comparison to the *in vitro* polarization. The effect of AA on monocyte polarization was evaluated. Finally, monocytes were analyzed for cytokine production of TNF and IL-8 by intracellular staining.

Both LPS and LTA induced polarization in healthy monocytes *in vitro*, with increased expression of both pro- (CD40 and PDL1, p<0.05) and anti-inflammatory (CD16 and CD163, p<0.05) polarization markers. This pattern resembled that of monocytes from septic patients. Treatment with AA significantly inhibited surface expression of CD16 and CD163 (p<0.05) in a dose dependent manner. Finally, AA attenuated LPS or LTA-induced cytokine production of IL-8 and TNF (both p<0.05) in a dose-dependent manner.

Thus, AA attenuates cytokine production and upregulation of anti-, but not pro-inflammatory related markers in LPS or LTA polarized monocytes. This study provides important insight into the effects of high dose AA on monocytes, and potential implications in sepsis.

**Summary sentence:** Ascorbic acid inhibits production of IL-8, TNF, and upregulation of the polarization markers CD16 and CD163 in LPS or LTA polarized monocytes *in vitro*.

## INTRODUCTION

Sepsis occurs due to a dysregulated immune response to infection. Current treatments include anti-infective therapy and supportive care, but no treatment targeting the dysregulated immune system is in clinical use. High dose ascorbic acid (AA) / Vitamin C has emerged as a potential treatment and has showed early promising results (1, 2). Additionally, AA has displayed anti-inflammatory and anti-oxidative effects in several animal models of sepsis (3-6). However, recent randomized controlled studies have failed to demonstrate a general benefit of AA treatment in sepsis and septic shock (7-10). Though, these studies did not thoroughly distinguish among different types of sepsis, pathogens or immunological states.

Few studies have addressed the effects of AA on the immune response in sepsis. Monocytes are immune cells with roles in cytokine production, antigen presentation and phagocytosis. In sepsis, monocytes are highly activated, or polarized, expressing markers related to scavenging (CD163), phagocytosis (CD16) and antigen presentation (CD40, PDL-1)(11-13). Still, it is important to note that conflicting data exists regarding surface markers in sepsis, likely reflect timing and subgroups of sepsis. Functionally, septic monocytes display a state of exhaustion with immunosuppressive features, increased ROS generation and intact or increased phagocytosis (14).

To the best of our knowledge, there has been no studies investigating the effects of AA on monocyte polarization. Here, we aimed to investigate the effects of high dose AA on monocyte polarization and function *in vitro* within the setting of sepsis.

## MATERIALS AND METHODS

### Patients

Healthy controls (n=6) and patients with blood-culture confirmed sepsis (n=3) were included in this study. Ethical approval was obtained from the Swedish Ethical Review Authority (2019-05146). The clinical characteristics of the septic patients have previously been described in *Kahn et. al*. (15).

### Surface marker expression in whole blood from patients with sepsis

Blood was collected in EDTA tubes from healthy controls (n=3) or sepsis patients (n=3) upon informed consent. 100μl of EDTA blood was stained with lineage markers and anti-CD40, PDL1, CD16 and CD163 (see **supplementary materials and methods** for details). Analysis was performed by a CytoFLEX flow cytometer (Beckman Coulter).

### Monocyte isolation and culture

Blood was collected in heparin tubes from healthy controls. A density gradient (LymphoPrep, Alere technologies) was used to collect the peripheral blood mononuclear cells (PBMCs). The cells were washed once with PBS and monocytes were further isolated through CD14 positive selection with magnetic beads using the MACS LS columns according to the manufacturer’s instructions (Miltenyi Biotech). 0.25×10^6^ cells were cultured in 500μl of RPMI-1640 medium with 2.05mM L-Glutamine (Gibco Life Technologies) supplemented with 10% normal human serum (Sigma-Aldrich). The monocytes were cultured at 37°C, 5% CO_2_.

L-Ascorbic acid (AA, Sigma-Aldrich) was prepared fresh by dilution in medium prior to each experiment. The possible effects of AA induced toxicity and pH changes were investigated and were considered minor (See **Supplementary Materials and Methods** and **Supplementary Figure 1A-B**). 125µg/ml of AA was used for experiments if not otherwise indicated.

### Polarization and flow cytometry

Monocytes from healthy controls (n=6) were polarized *in vitro* by addition of LPS (10ng/ml, Invivogen) or LTA (1μg/ml, Sigma-Aldrich), simulating Gram-negative and Gram-positive polarization, respectively, and subsequently cultured for 48hrs, with or without AA. In some experiments, AA was instead added after 24hrs of stimulation with LPS or LTA. Next, cells were detached with PBS/0.5mM EDTA and gentle pipetting. The monocytes were washed and resuspended in 50μl PBS. The monocytes were subsequently stained for anti-CD40, PDL1, CD16 and CD163 (see **Supplementary Materials and Methods** for details). The cells were analyzed by the CytoFLEX.

### Intracellular cytokine production

Monocytes from healthy controls were isolated as described above. Following isolation, medium was supplemented with 1μl/ml of BD GolgiPlug (BD Biosciences) and 1ng/ml LPS or 100ng/ml LTA as inducers of cytokine production. When used, AA was added directly and monocytes were cultured for 5hrs before detachment using PBS/0.5mM EDTA and fixated/permeabilized using the BD CytoFix/Perm kit (BD Biosciences) according to the manufacturer’s instructions. The monocytes were subsequently stained for anti-TNF (clone: MAb11, PE) and anti-IL-8 (clone: E8N1, Alexa fluor 488), all from BD, diluted 1:50, for 30min at 4°C and analyzed by a CytoFLEX.

### Statistics

Statistical analysis was performed using Prism 8. Values are presented as median if not otherwise indicated. Paired data was analyzed using the Wilcoxon signed ranked pairs test. p<0.05 was considered statistically significant.

## RESULTS AND DISCUSSION

Septic monocytes have previously been shown to be activated as evidenced by increased expression of polarization markers such as CD16, CD163, PDL-1 and CD40, amongst others (11-13). However, results on expression of polarization markers are conflicting, likely reflecting timing and different phenotypes amongst patients. Thus, we sought to confirm previously described changes in monocyte polarization in our setting. Monocytes in whole blood from septic patients (n=3) display signs of polarization, i.e., increased expression of CD16, CD40, CD163 and PDL-1 compared to controls (n=3) (**Figure 1A-B**). Next, we sought to explore the influence of two bacterial components, lipopolysaccharide (LPS) and lipoteichoic acid (LTA), on polarization of healthy monocytes. Stimulation of healthy monocytes (n=6) resulted in significant upregulation of CD16, CD40, CD163 and PDL-1 (p<0.05) upon 48hrs of LPS stimulation (**Figure 2A-B**). Histograms of a representative LPS stimulated sample can be found in **Figure 2A**. CD16, CD163 and PDL1 were upregulated following polarization with LTA (p<0.05) (**Figure 2C**). Thus, polarization could be induced by LPS or LTA and this polarization pattern resembles, at least partly, that of monocytes from patients with sepsis.

**Figure 1.**
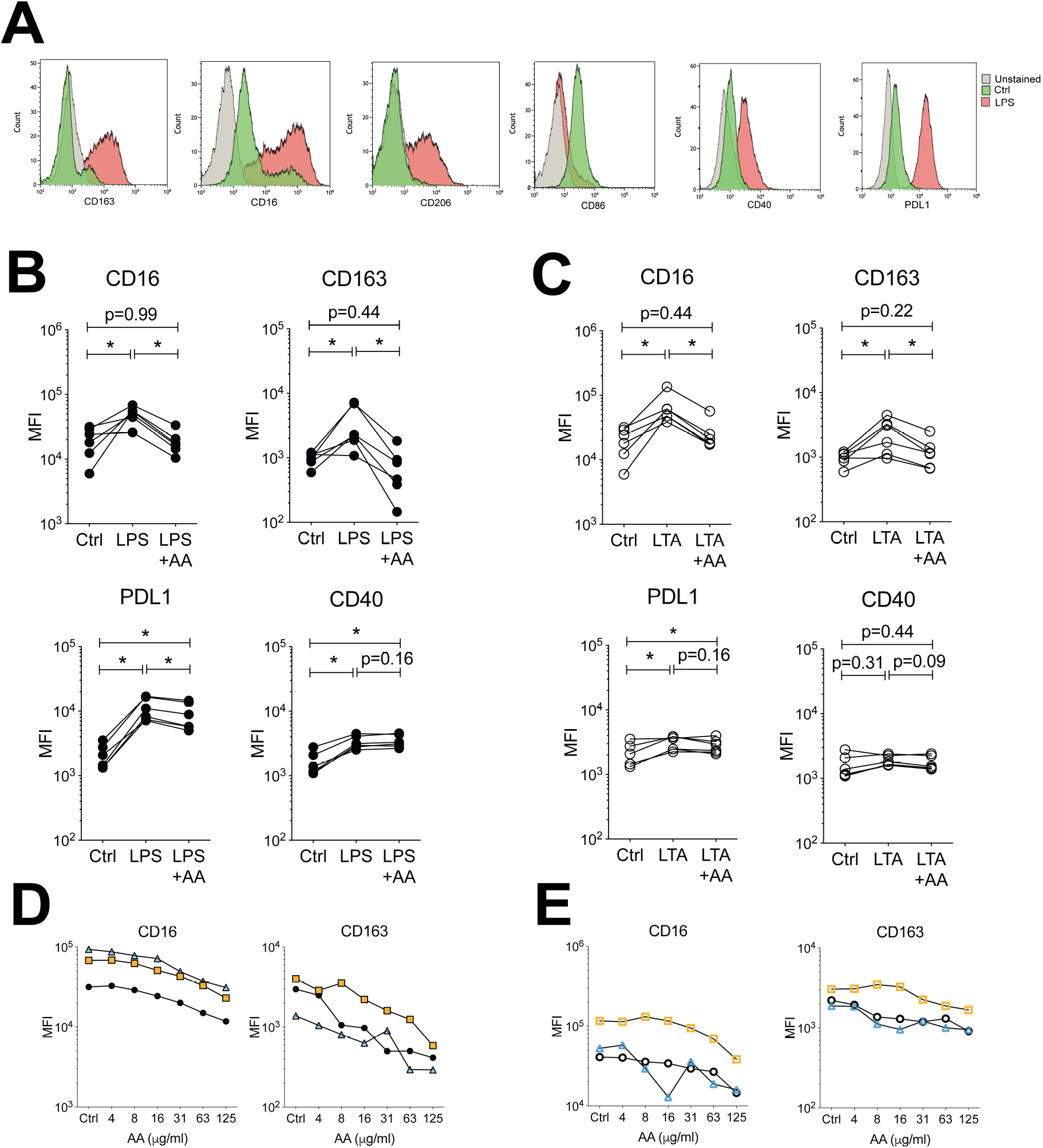
Expression of polarization markers in patients with sepsis. Monocytes in whole blood from sepsis patients (n=3) or healthy controls (n=3) were analyzed for the expression of four polarization markers. (**A**) shows a representative gating strategy in a patient with sepsis. (**B**) shows the expression of polarization markers in sepsis or healthy controls. Line at median

**Figure 2.**
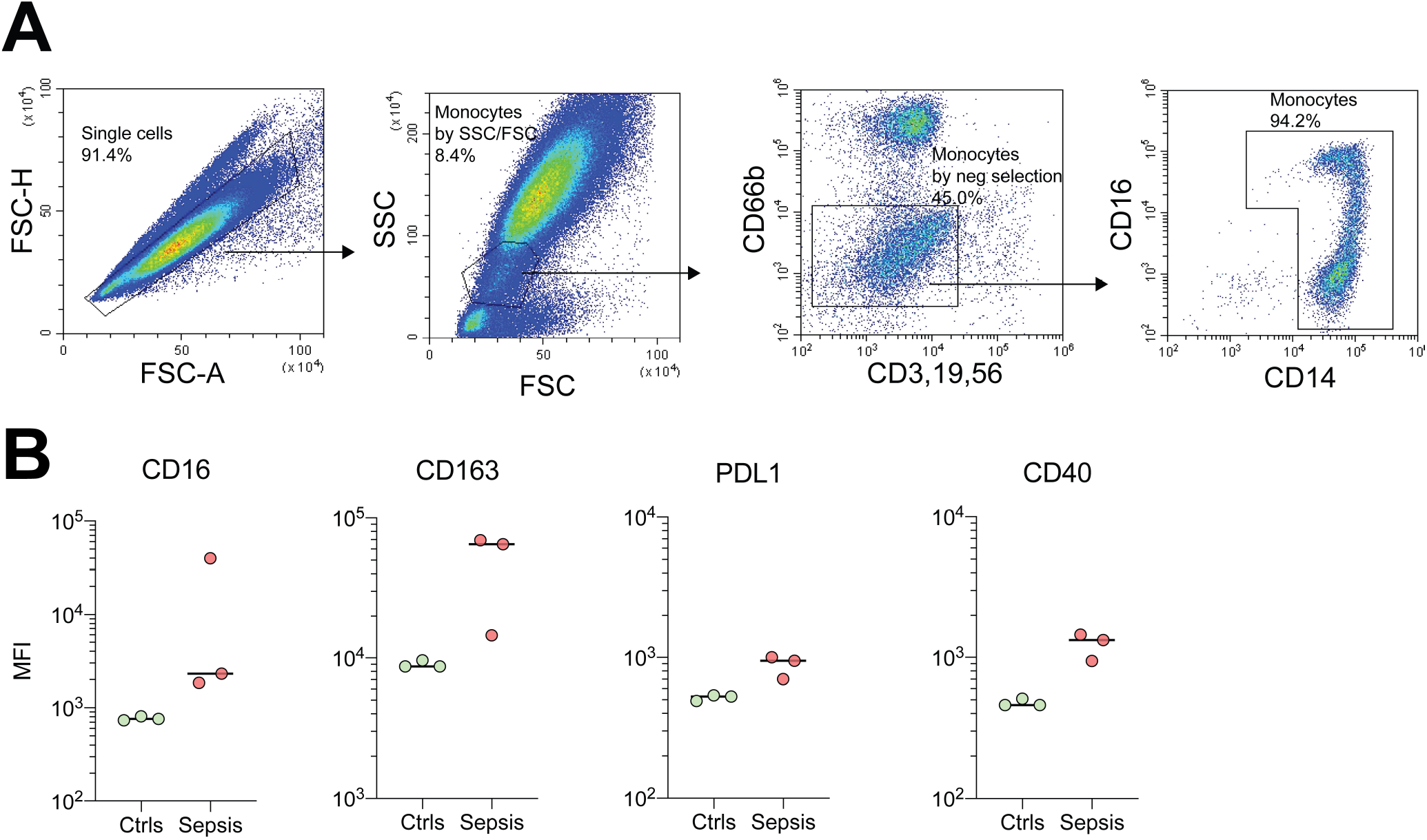
Ascorbic acid inhibits upregulation of CD16 and CD163, but not CD40 and PDL-1, in LPS or LTA polarized monocytes *in vitro*. Monocytes were isolated from healthy controls (n=6) and polarized for 48hrs or not (ctrl) before analysis of surface markers by flow cytometry. In addition, monocytes were treated or not with 125µg/ml of AA. (**A**) shows representative histograms of the four markers of one LPS induced control. (**B**) Shows LPS polarized monocytes with or without AA treatment and (**C**) similarly shows LTA polarized monocytes. The effects of AA were dose-dependent in n=3 controls polarized with (**D**) LPS or (**E**) LTA. Data is presented as median fluorescence intensity (MFI). Statistics were performed using the Wilcoxon matched-pair signed rank test. *p<0.05. *LPS – Lipopolysaccharide, LTA – lipoteichoic acid, AA – Ascorbic acid*

Ascorbic acid (AA) has been suggested as treatment for multiple diseases, ranging from cancer to sepsis. Patients with sepsis have low levels of AA, which can be reversed upon AA treatment (1). The general mechanisms of AA in sepsis are believed to be anti-inflammatory and anti-oxidative (16). Multiple animal and disease models of sepsis have shown benefits of AA treatment on several organ systems, e.g., the vascular and pulmonary systems (17-20). However, recently, several randomized control trials failed to demonstrate a general beneficial effect of AA in sepsis patients (7-10). Although, these studies did not thoroughly distinguish among different types of sepsis, pathogens, or immunological states. Hence, AA might have potential in specific windows-of-time or in certain subgroups.

Few studies have investigated effects of AA treatment on the immune response in sepsis. AA has been shown to restore several effector functions of septic neutrophils *ex vivo*, e.g., phagocytosis and NETosis (21). In monocytes, dehydroascorbic acid was shown to protect against LPS induced oxidative stress (22). AA has also been shown to inhibit ROS production, NFkB activation and to inhibit apoptosis (23-25). Thus, AA influences multiple aspects of the immune system, but we now little of its mechanisms in sepsis.

To investigate if AA could modulate monocyte polarization, monocytes were stimulated with LPS or LTA, with and without supplemented AA. AA at 125µg/ml attenuated the expression of CD16 and CD163 in LPS or LTA polarized monocytes to that of the controls (p<0.05, **Figure 2B-C**) and this inhibition was dose dependent (**Figure 2D-E)**. In LPS polarized monocytes, AA slightly reduced the expression of PDL-1, but not CD40 (**Figure 2B**). Changes in the expression of PDL-1 and CD40 in LTA polarized monocytes were minor (**Figure 2C**). AA did not have a significant effect on the markers in unstimulated monocytes (**Supplementary Figure 2**). Thus, AA attenuates the expression of CD16 and CD163, but not CD40 and PDL-1, in LPS and LTA polarized monocytes.

Traditionally, CD16 and CD163 are markers of anti-inflammation, or M2, polarization and are upregulated by various stimulus *in vitro*, such as IL-10. These monocytes, or macrophages, have traditionally been termed suppressive. However, it is important to note the complexity of polarization. CD163^+^ monocytes produce more pro-inflammatory cytokines than CD163^-^ monocytes in sepsis (14). In addition, monocytes display a suppressive and exhausted phenotype and when exposed to LPS they respond poorly with production of pro-inflammatory cytokines, as opposed to monocytes from patients with hyperinflammatory COVID-19 (15). Thus, these data suggest that AA might not target only the pro-inflammatory aspect (inflammatory cytokine production) but also the immunosuppressive phenotype (CD16 and CD163) of monocytes in sepsis.

Next, we investigated if the effects of AA are due to a downregulatory or inhibitory mechanism. Addition of AA to monocytes 24hrs after LPS or LTA stimulation, instead of at the start of culture, did not result in altered expression of CD16 and CD163 compared to monocytes not receiving AA (**Figure 3A-B**). Thus, AA inhibits upregulation-, rather than downregulates, the observed surface markers. This observation is important in the aspect of future study designs. This data highlights the role of timing of AA treatment, and could be a potential factor contributing to the lack of benefit in the conducted studies.

**Figure 3.**
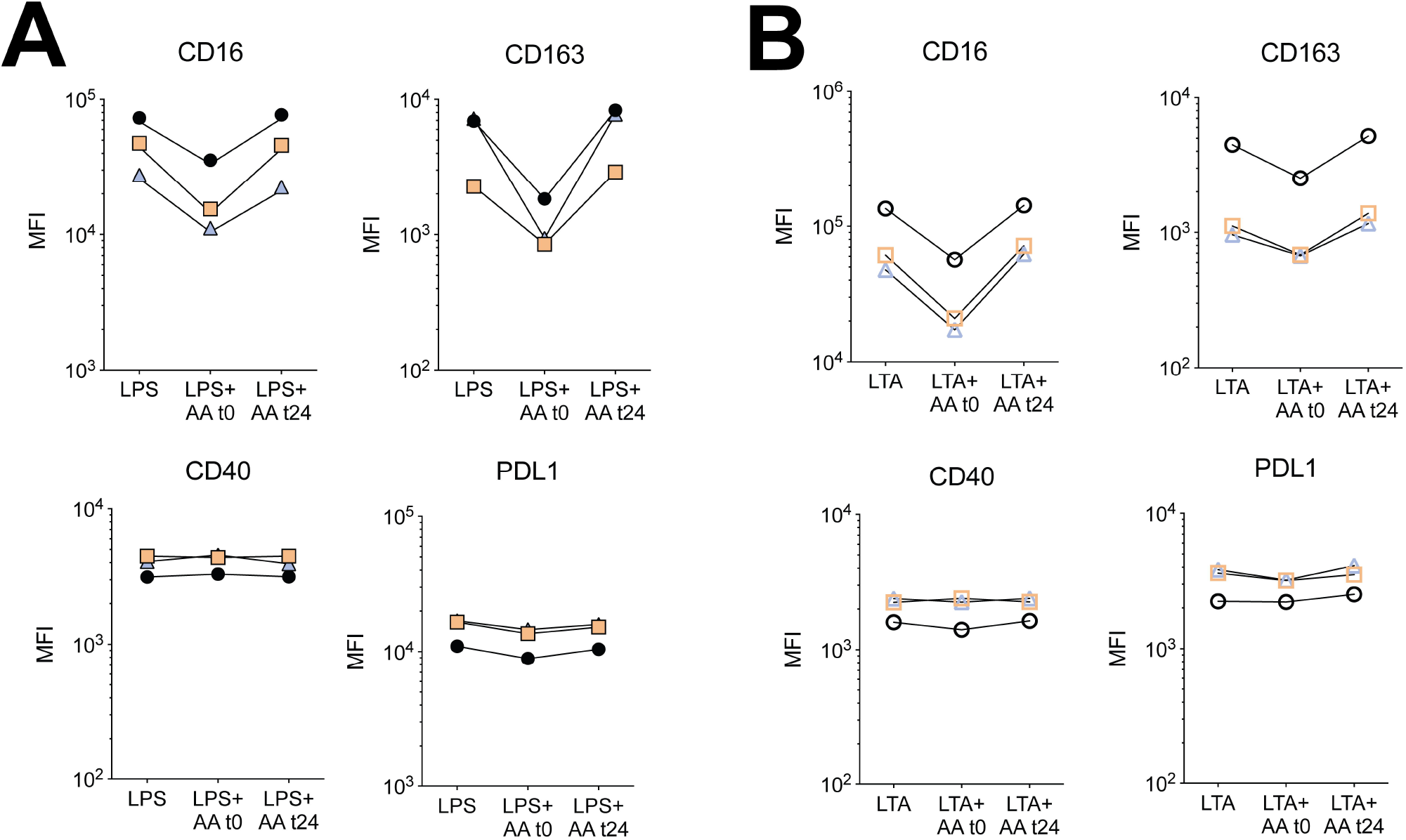
ascorbic acid inhibits upregulation, rather than downregulates, the surface markers induced by LPS or LTA. Monocytes were isolated from healthy controls (n=3) and polarized for a total of 48hrs with (**A**) LPS or (**B**) LTA and analyzed for the expression of polarization markers. At the time of polarization (t0) or after 24hrs of culture (t24) AA was added at 125µg/ml. Data is presented as median fluorescence intensity (MFI). *LPS – Lipopolysaccharide, LTA – lipoteichoic acid, AA – Ascorbic acid*

To investigate whether AA affected monocyte function, we assessed cytokine production of IL-8 and TNF in healthy monocytes (n=6). Both LPS and LTA induced production of IL-8 and TNF which was attenuated by AA (p<0.05) (**Figure 4A-C**). This effect was also dose dependent (**Figure 4D**). **Figure 4C** shows representative histograms of LTA activation in one control. Thus, AA affects monocytes on the functional level by inhibiting production of IL-8 and TNF. Using a similar method, a previous study found that AA attenuates LPS induced production of IL-6 and TNF, but not IL-8 in monocytes in whole blood (26). The use of whole blood, rather than purified monocytes, could be a major contribute to the discrepancy between our results. Nevertheless, we further confirm that AA influences monocytes function by inhibiting pro-inflammatory cytokine production.

**Figure 4.**
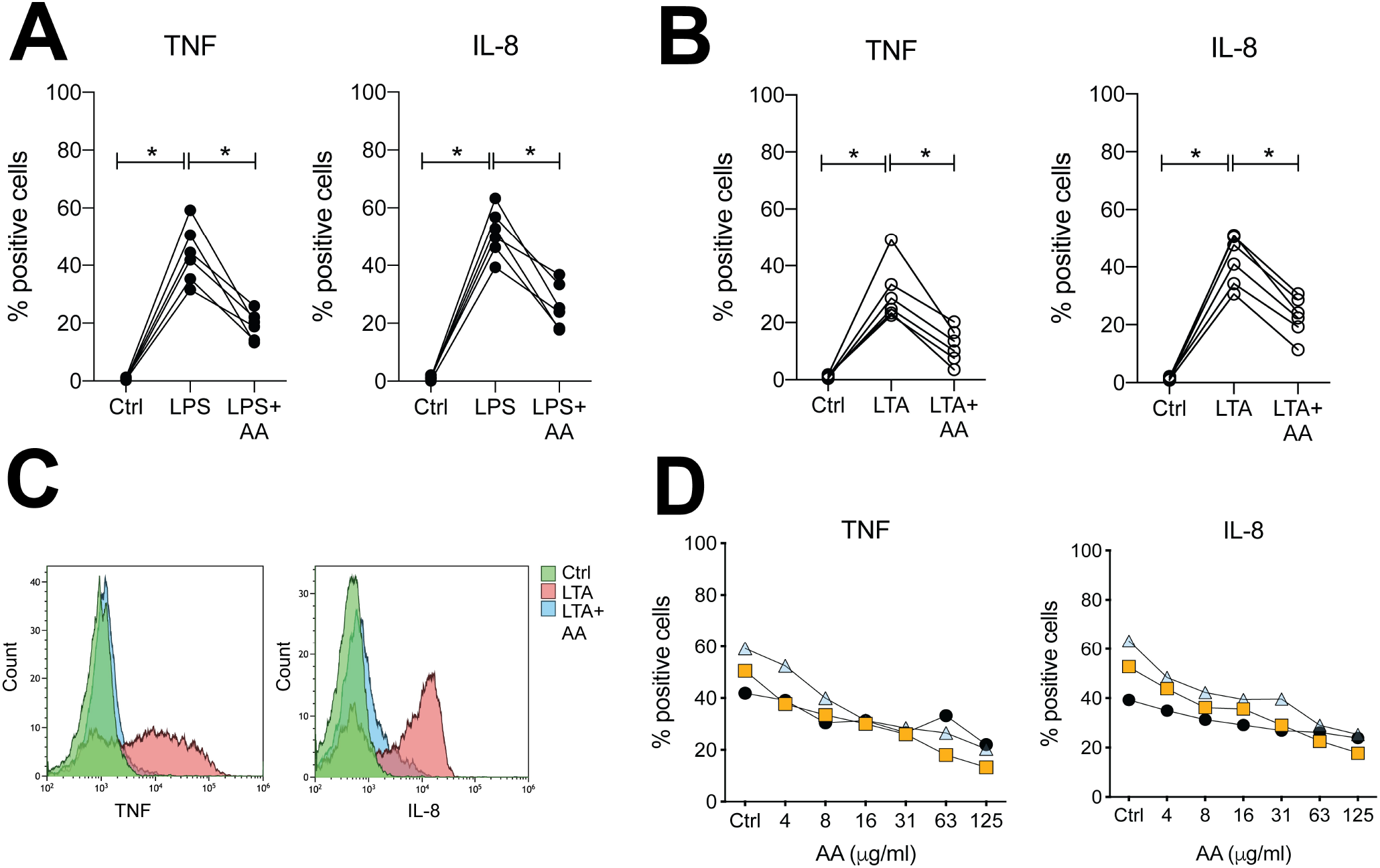
Ascorbic acid attenuated LPS or LTA induced cytokine production. Monocytes were isolated from healthy controls (n=6) and treated with brefeldin A followed by LPS or LTA, with or without AA treatment. The cells were cultured for 5hrs and analyzed for intracellular accumulation of TNF or IL-8. Both (**A**) LPS and (**B**) LTA significantly induced production of TNF and IL-8 in monocytes, which was attenuated by treatment with AA. (**C**) shows representative histograms of one control stimulated with LTA. The effects of AA on LPS (**D**) and LTA (**E**) polarized monocytes were dose-dependent as observed in n=3 controls Statistics were performed using the Wilcoxon matched-pair signed rank test. *p<0.05. *LPS – Lipopolysaccharide, LTA – lipoteichoic acid, AA – Ascorbic acid*

The use of LPS and LTA polarized monocytes as opposed to monocytes from patients with sepsis constitutes a limitation in this study. Even though LPS and LTA polarized monocytes display similarities with septic monocytes, the use of these cells would be important to explore in future studies. In addition, more functional studies need to be performed to investigate the influence of AA on monocytes, and their influence with the surrounding environment.

In conclusion, we show that high concentrations of AA attenuate LPS or LTA induced surface expression of CD16 and CD163, but not CD40 and PDL-1. Furthermore, AA inhibits LPS or LTA induced cytokine production of TNF and IL-8. These data provide an important insight into the effect of AA on monocytes in relation to sepsis.

## Data Availability

Data is available from the corresponding author upon reasonable request

## AUTHORSHIP

T.S. carried out the experiments, T.S., R.K. and F.K. interpreted data and wrote the manuscript.

F.K. conceptualized and designed the study and collected patient data. All authors have approved the final manuscript, critically revised it and agreed to be accountable for all aspects of the work.

## ACKNOWLEDGMENTS

Dr. Anki Mossberg and Dr. Birgitta Gullstrand are humbly acknowledged for their aid in handling of patient samples. We thank Sabine Arve-Butler, Olivia Aherne, Anki Mossberg and Louise Thelaus for their critical input on the manuscript. Robin Kahn is currently receiving grants from the Swedish Rheumatism Association, Greta and Johan Kock’s Foundation, the Anna-Greta Crafoord Foundation, the Crafoord Foundation, the Swedish Medical Society, Alfred Österlunds Foundation, The Knut and Alice Wallenberg foundation, the Medical Faculty at Lund University and Region Skåne. Fredrik Kahn is currently receiving grants from the Crafoord Foundation, Alfred Österlunds Foundation, the Swedish Research council, the Medical Faculty at Lund University and Region Skåne.

## CONLICT OF INTEREST DISCLOSURE

The authors declare no conflict of interest

## Abbreviation page

*AA*: *Ascorbic acid*
*MFI*: *Median Fluorescence Intensity*
*LPS*: *Lipopolysaccharide*
*LTA*: *lipoteichoic acid*
PBMC: *Peripheral blood mononuclear cells*

