## Supplementary Materials and Methods for "Ascorbic acid attenuates activation and cytokine production in sepsis-like monocytes"

**Surface marker expression in whole blood**

Antibodies used to stain monocytes in whole blood in sepsis patients and controls: anti-CD16 (clone: 3G8, PerCP-Cy5.5), anti-CD163 (clone: GHI/61, PE-CF594), anti-CD66b (clone: G10F5, Alexa fluor 647), anti-CD14 (clone: M5E2, Alexa fluor 700), anti-CD206 (clone: 15-2, APC/Fire750), anti-PDL1 (clone: MIH1, Bv421), anti-CD40 (clone: 5C3, Bv510), anti-CD86 (clone: FUN-1, BV650), anti-CD19 (clone: HIB19, Bv786), anti-CD3 (clone: UCHT1, Bv786) and anti-CD56 (clone: NCAM16.2, Bv786) in brilliant violet staining buffer (BD Biosciences). All BD Biosciences except CD14, CD16 and CD206 (Biolegend). The cells were incubated in dark at room temperature (RT) for 30min. Next, red blood cells were lysed using lysis solution (BD Biosciences) and remaining cells were washed twice with PBS. Finally, cells were resuspended in PBS before analysis.

**Toxicity and pH changes of ascorbic acid**

Possible effects on viability was assessed with Annexin V and propidium iodide staining (BD Biosciences) according to the manufacturer’s instructions. Cells were viable in the range of 4-125µg/ml of vitamin C, and subsequently started to lose viability (**Supplementary Figure 1A**). The effect of pH was investigated using NaHCO_3_ as a buffer, and the pH was measured using a pH meter (Mettler Toledo). In some experiments, HCl was used to lower pH. AA reduced pH slightly from 7.6 to 7.2, but this pH change did not influence the effects of AA (**Supplementary Figure 1B**).

**Staining of polarization markers in cultured monocytes**

The following markers were used to stain cultured monocytes: anti-CD40 (clone: 5C3, BV510), anti-CD274/PDL-1 (clone: MIH1, BV421) anti-CD16 (clone: 3G8, APCH7), anti-CD86 (clone: FUN-1, FITC), anti-CD206 (clone: 19.2, APC) and anti-CD163 (clone: GHI/61, PE), all from BD. The antibodies were diluted 1:50 in FACS buffer (PBS with 0.5% BSA) in 100μl. The cells were stained for 30min, RT, washed once and resuspended in 150μl PBS for analysis by flow cytometry (CytoFLEX, Beckman Coulter).

**FIGURE LEGENDS**

**Supplementary Figure 1 The effect of ascorbic acid on cell viability and pH** (**A**) Monocytes were isolated from healthy controls (n=3) and polarized with LPS and treated with AA at different concentrations for 48hrs. The control well received no AA treatment. Viability of monocytes was assessed in a dose dependent manner and analyzed using annexin V and propidium iodine (n=3) (**B**) AA at 125µg/ml slightly reduced the pH from 7.6 to 7.2 in medium. Restoration of pH was performed using NaHCO_3_ as a buffer. In parallel, HCl was used to lower to pH of medium to 7.2 to mimic the effect of AA on pH. Monocytes were cultured for 48hrs and analyzed for expression of six activation markers by flow cytometry. Line at median. *LPS – Lipopolysaccharide, AA – Ascorbic acid*

**Supplementary Figure 2 The effect of ascorbic acid on healthy unpolarized monocytes** Monocytes were isolated from healthy controls (n=6) and treated with AA or not for 48hrs before analysis by flow cytometry for expression of six activation markers. Data is presented as median fluorescence intensity (MFI). Statistics were performed using the Wilcoxon matched-pair signed rank test.
