## Supplementary figures and images for "Ascorbic acid attenuates activation and cytokine production in sepsis-like monocytes"

### Supplementary figure 1

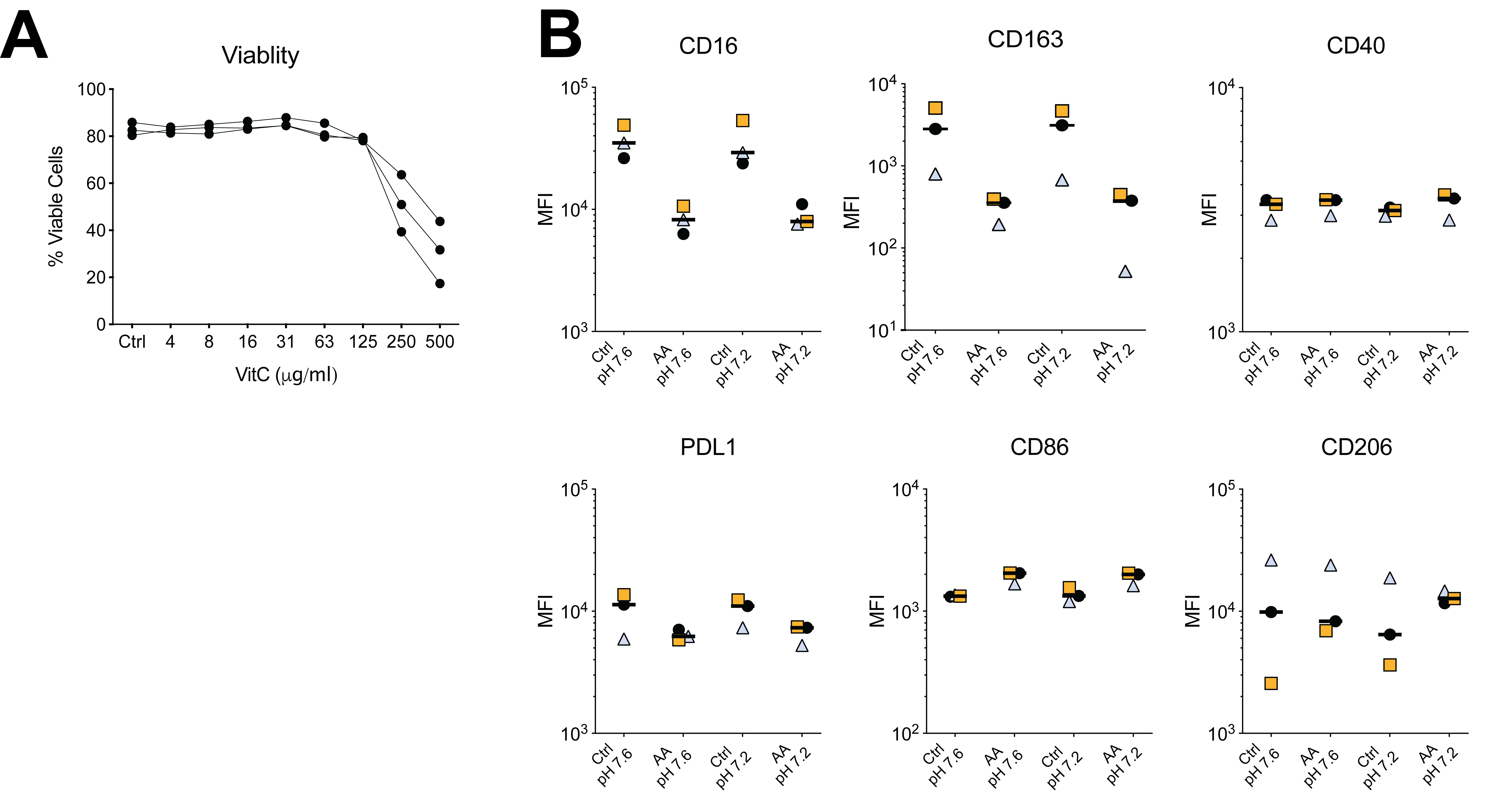

### Supplementary figure 2

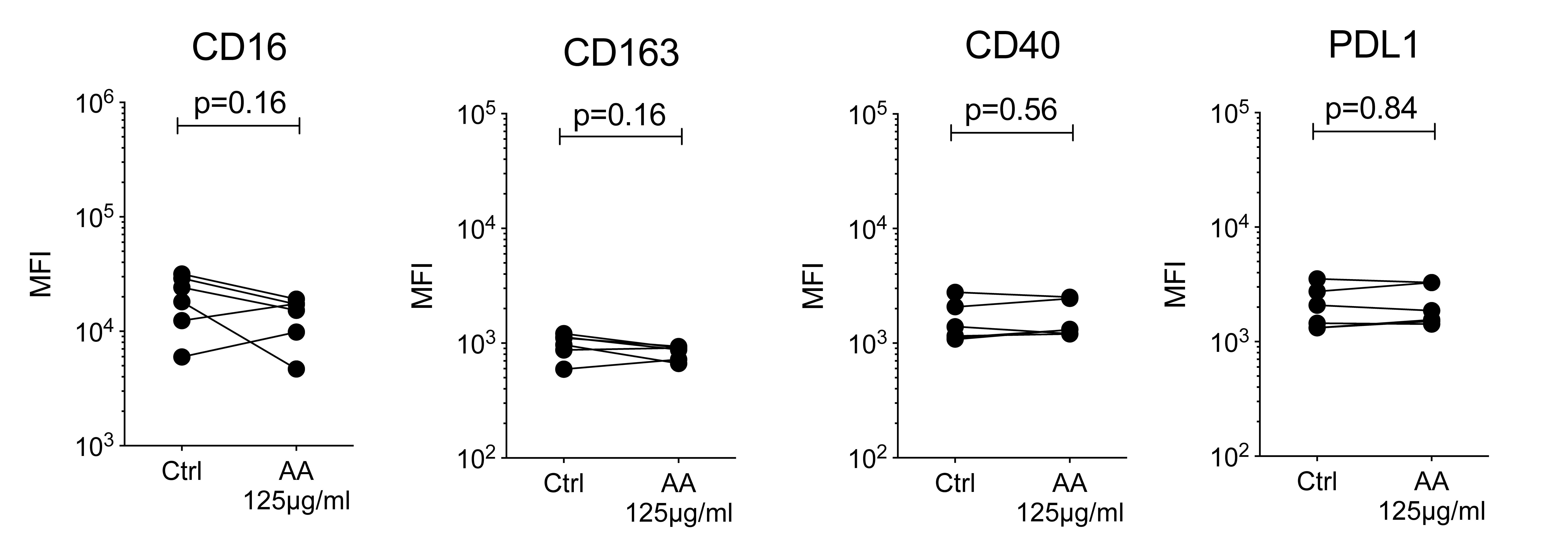
